# A scoping review of behavioural science approaches and frameworks for health protection and emergency response

**DOI:** 10.1101/2023.05.19.23290226

**Authors:** Alice Zelenka Martin, Dale Weston, Jo Kesten, Clare French

## Abstract

**Aim:** Rapid intervention development, implementation and evaluation is required for emergency public health contexts, such as the recent COVID-19 pandemic. A novel Agile Co-production and Evaluation (ACE) framework has been developed to assist this endeavour in future public health emergencies. This scoping review aimed to map available behavioural science resources that can be used to develop and evaluate public health guidance, messaging, and interventions in emergency contexts onto components of ACE: rapid development and implementation, co-production with patients or the public including seldom heard voices from diverse communities, and inclusion of evaluation.

**Methods:** A scoping review methodology was used. Searches were run on MEDLINE, EMBASE, PsychInfo, and Google, with search terms covering emergency response and behavioural science. Papers published since 2014 and which discussed a framework or guidance for using behavioural science in response to a public health emergency, were included. A narrative synthesis was conducted.

**Results:** Seventeen records were included in the synthesis. The records covered a range of emergency contexts, the most frequent of which were COVID-19 (n=7) and non-specific emergencies (n=4). One record evaluated existing tools, six proposed new tools, and ten described existing tools. Commonly used tools included the Behavioural Change Wheel, Capability, Opportunity, and Motivation Behaviour model (COM-B model) and social identity theory. Three records discuss co-production with the target audience and consideration of diverse populations. Four records incorporate rapid testing, evaluation, or validation methods. Six records state that their tool is designed to be implemented rapidly. No records cover all components of ACE.

**Conclusion:** We recommend that future research explores how to create guidance involving rapid implementation, co-production with patients or the public including seldom-heard voices from diverse communities, and evaluation.

## Introduction

The role of health-protective human behaviour in combatting the spread of COVID-19 has highlighted the importance of behavioural science as a vital tool in the health protection and emergency response arsenal.^1^ Guidance for utilising behavioural science in routine public health is available.^2-4^ However, the challenge of developing and utilising behavioural science in a public health emergency and evaluating the implementation of interventions is that appropriate public health response behaviours need to be understood, and guidance must be formulated and disseminated rapidly to be implemented in time to prevent avoidable outcomes.^5, 6^ It also needs to be responsive to evolving situations and able to be updated as the evidence-base changes.^7, 8^ For guidance to be effective, it needs to be delivered by a trusted source, consistent across all responding organisations, be credible and realistic.^5, 6, 8-10^ These standards can be challenging to meet under time pressure, especially when key behavioural science frameworks are typically designed for non-emergency health contexts.^4, 11, 12^ There is therefore a clear need to establish an evidence-based, effective framework for conducting rapid, emergency preparedness and response-focused guidance and intervention development, ahead of any future public health emergencies.

The ACE (Agile Co-production and Evaluation) framework aims to fulfil this need by supporting the rapid development and evaluation of public health interventions, messaging and guidance involving co-production with target populations.^13^ This framework focuses on 1) a need to rapidly develop and disseminate messages, guidance or interventions; 2) consideration of how to involve and engage members of the public, especially under-served communities whose voices are seldom heard or incorporated into intervention development, and 3) use of rapid testing, evaluation or validation methods (such as online testing or implementation evaluation using routine data) to provide feedback on effectiveness. These three points have been identified by the authors of this review (and co-authors of the ACE framework) as central to the approach; these criteria will be referred to hereafter as the ACE criteria.

Although the authors are unaware of any existing emergency preparedness and response focused behavioural science framework that addresses all of these constructs, this scoping review aimed to map the extent and type of available resources concerning behavioural science approaches and frameworks for health protection in emergency contexts. We wanted to gain insights into the extent to which existing tools have been designed for rapid implementation and co-production with the target population. This will help to inform the ACE framework by leading to the development of recommendations for future work, with the aim of creating a framework for effective and timely implementation of interventions, guidance and messaging as part of a public health emergency response.^14^

## Methods

The Joanna Briggs Institute (JBI) scoping review framework was used.^15^ A review protocol, based on the JBI template,^16^ is published on the Open Science Framework. DOI: 10.17605/OSF.IO/FE6TN.

The literature searches were conducted on MEDLINE, EMBASE, and PsychINFO on the 16^th^ and on Google on the 17^th^ of June 2022 by AZM. The search terms (Supplementary Material 1) covered two core topics: behavioural science and emergency response. We limited the search to records published since 2014; this was a pivotal year for the incorporation of behavioural science into emergency response^17^ and a crucial time in work to standardise and collate theories and approaches in behavioural science.^18, 19^ A Google search captured grey literature, such as government and other public health-related documents (Supplementary Material 1). Only the first ten pages of Google results ordered by relevance were screened.

The records, including the grey literature, were title and abstract, and full text screened in duplicate on Rayyan (https://www.rayyan.ai/) using our pre-specified inclusion and exclusion criteria. AZM screened all records with duplicate screening of each record conducted by DW, JK or CF. Conflicting decisions between reviewers were resolved through a whole-group discussion.

Inclusion criteria:

i. Type of source: any type of record is eligible.
ii. Subject: the record must address guidance or frameworks for utilising behavioural science in an emergency response context; this may include evaluations or assessments of existing guidance/frameworks, discussions on developing new guidance/frameworks, and discussions on the presentation of existing guidance/frameworks.
iii. Context: behavioural science must be applied to an emergency of public health relevance. Records published or covering emergencies in any country are eligible.
iv. Other: records published since 2014. English or with an English version available.

AZM performed data extraction using a pre-defined Excel database, and co-authors discussed the results. Data were extracted based on JBI guidance^15, 16^ and the ACE criteria.^13^ Data were synthesised to identify: a) who is producing guidance and frameworks; b) emergency contexts; c) co-production with patients or the public; d) consideration of how to engage diverse populations; e) to what extent tools incorporate testing or evaluation; and, f) who tools are being written for. Groupings a to e were pre-specified, and grouping f developed iteratively as we synthesised the data. Results were narratively synthesised.

## Results

The database and Google searches identified 2,449 records. Following the removal of duplicates, 2,049 records underwent title and abstract screening. Sixty-two records were sought for full-text screening, but four could not be retrieved. Seventeen records were included for data extraction (Fig. 1).

**Figure 1.**
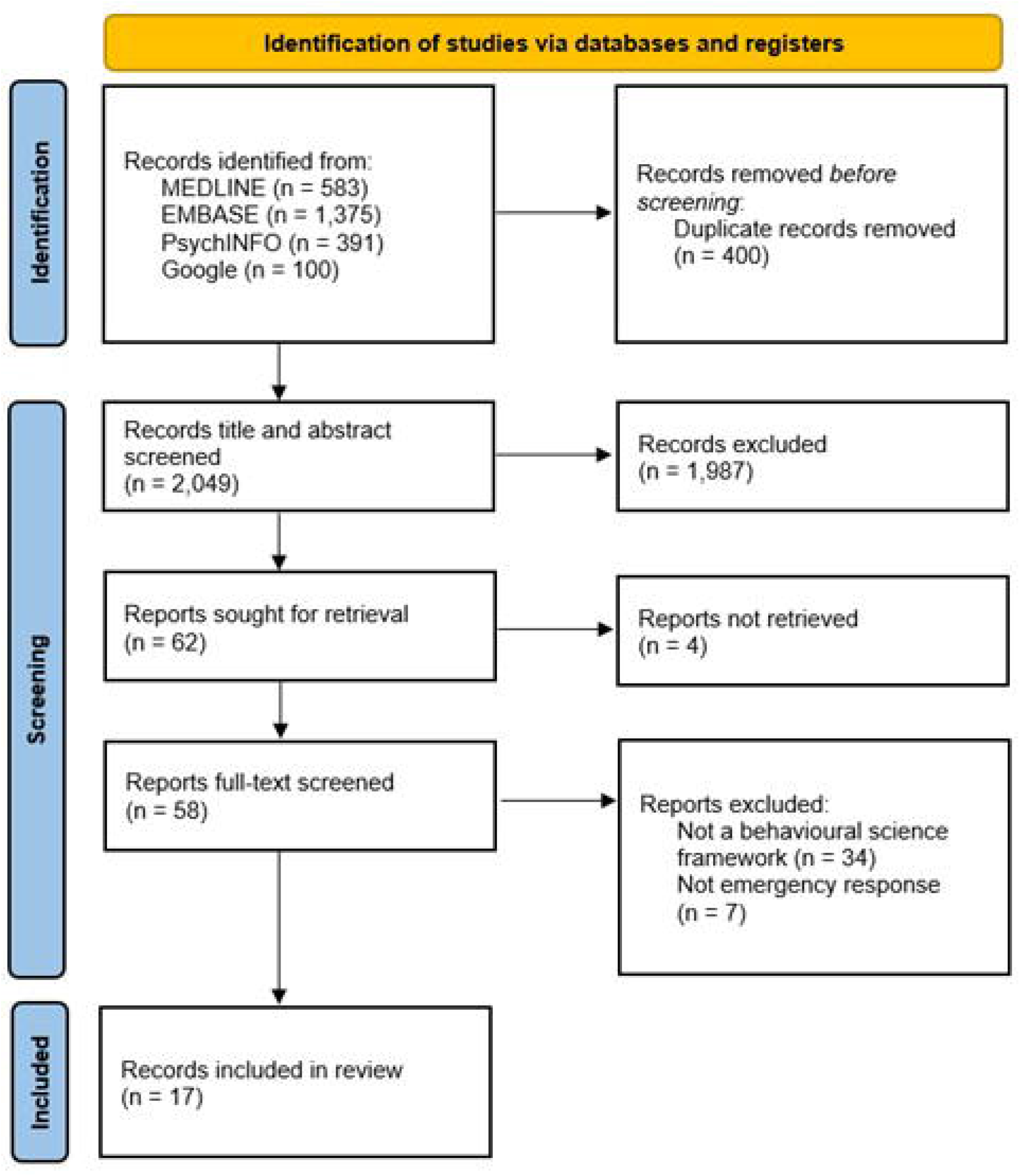

### Characteristics of included articles

Included studies were primarily peer-reviewed journal articles, but there was also a policy paper^20^ and digital educational material.^21^ Eleven records were authored by a multidisciplinary group, bringing a range of professional and academic knowledge to their work (Fig. 2).

**Figure 2.**
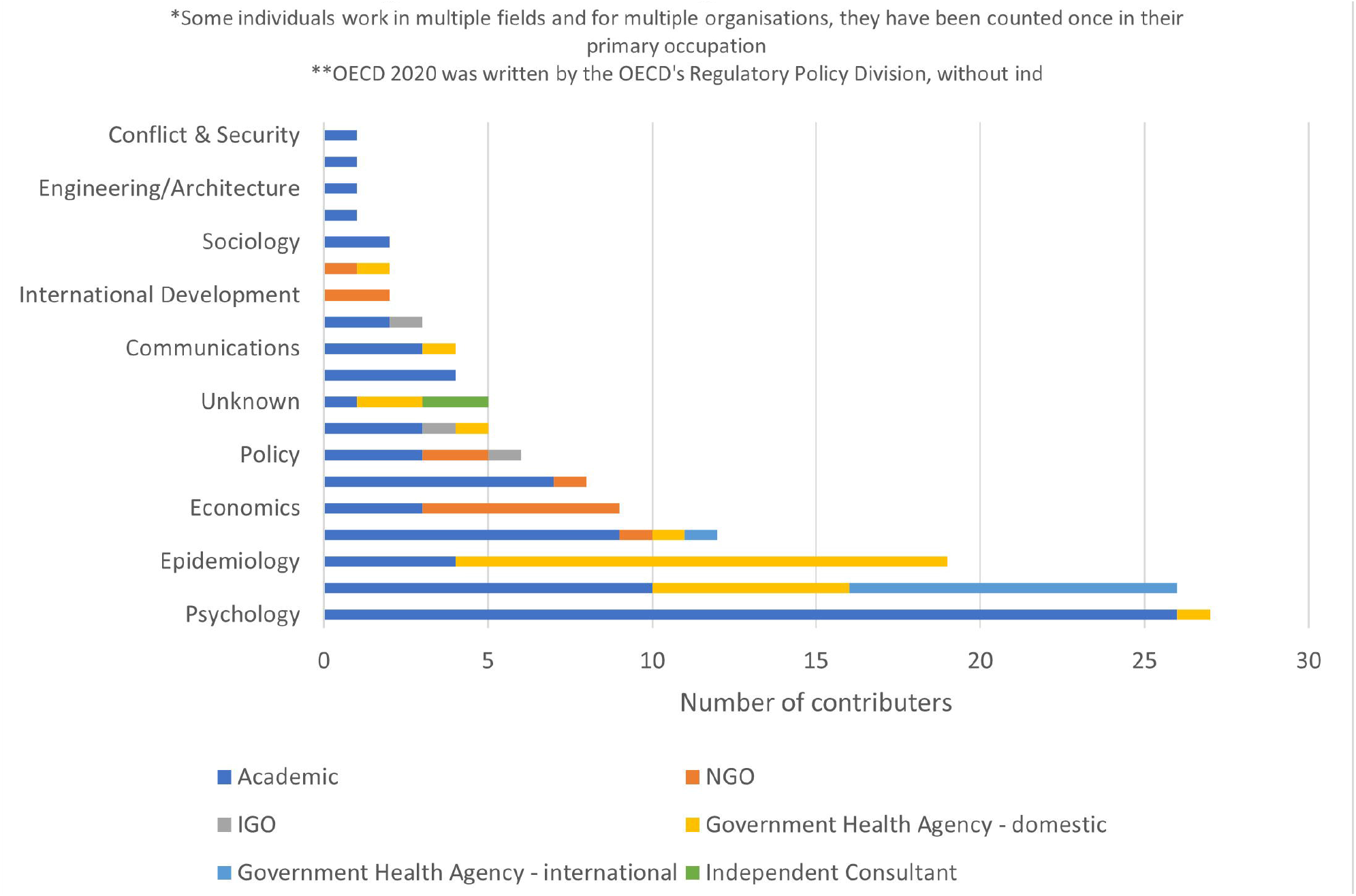
Professional Background of Contributors

Four studies were published prior to 2020.^22-25^ Seven records address the response to COVID-19, all published in 2020 and 2021.^5, 20, 26-30^ Other emergency contexts include flooding,^31^ natural disaster,^23^ chemical incident,^24^ Zika virus,^22^ non-specific infectious disease outbreak^32, 33^ and non-specific emergencies^7, 21, 25, 34^ (Table 1).

**Table 1:** Summary of included records.

|  | **Emergency context** | **Emergency response type** | **Behavioural Science theories & constructs** |
| --- | --- | --- | --- |
| **Jackson et al. 2017** | Natural Disasters | Health promotion interventions | Community mobilisation, social marketing |
| **Jalloh et al. 2021** | Infectious Disease Outbreak | Behaviour change implementation approaches | Top-down, intermediary & bottom-up approaches |
| **Lee et al. 2021** | COVID-19 | Behaviour change interventions | BCW |
| **Morton et al. 2021** | Non-specific | Adapting pre-existing interventions | Table of Changes method from the Person-Based Approach |
| **SBCC for Emergency Preparedness 2020** | Non-specific | Communication | Socio-ecological model |
| **West et al. 2020** | COVID-19 | Behaviour change interventions | Various models, but primarily COM-B, BCW & PRIME theory |
| **Kuhlicke et al. 2020** | Flooding | Personal protective behaviour | The 'Behavioural turn.' |
| **Michie et al. 2020** | COVID-19 | Increasing adherence to health protection behaviours | BCW |
| **Oxman et al. 2022** | Non-specific | Communication - persuading & informing | N/A |
| **Templeton et al. 2020** | COVID-19 | Increasing safe behaviour & adherence to guidance | Social identity theory |
| **Carter et al. 2015** | Chemical Incident | Crowd psychology | Social identity theory |
| **Chater et al. 2021** | COVID-19 | Rapid guidance development | TRICE, using BCW & COM-B |
| **OECD 2020** | COVID-19 | Applying behavioural insights to policy | ABCD & BASIC frameworks |
| **Pinchoff et al. 2019** | Zika Virus | Prioritising interventions | N/A |
| **Linnemayr et al. 2016** | Non-specific | Behavioural economics | Invoking social norms, changing the default, framing messages in terms of loss, plan-making & mental mapping, commitment devices, financial micro-incentives, representing small probabilities |
| **Lunn et al. 2020** | COVID-19 | Increasing adherence to personal protective behaviours | N/A |
| **Weston et al. 2020** | Infectious Disease Outbreak | Behaviour change theories | Health Belief Model, Theory of Planned Behaviour, & Protection Motivation Theory |

\*COM-B - Capability, Opportunity, and Motivation Model of Behaviour; BCW – Behaviour Change Wheel; TRICE - Template for rapid iterative consensus of experts; PRIME – Planning, Response, Impulse/Inhibition, Motive and Evaluation; ABCD - Attention, Belief formation, Choice and Determination; BASIC - Behavioural, Analysis, Strategy, Intervention and Change.

Four records specify a country where their emergency scenarios and response methods occurred.^5, 22, 28, 34^ The United Kingdom is the country most represented by the included studies.5, 28, 34

### Purpose & Audience

One record aimed to evaluate existing tools;^31^ six proposed new tools;^7, 24, 25, 29, 32, 34^ and ten described existing tools.^5, 20^-^23, 26^-^28, 30, 33^ The emergency response tools discussed predominantly use behaviour and behaviour change approaches,^5, 21, 23^-^28, 31, 33^ such as COM-B and Planning, Response, Impulse/Inhibition, Motive and Evaluation (PRIME) Theory, as well as implementation,^32^ decision making,^22, 34^ intervention development,^7, 20^ and communication theories and approaches.^29, 30^

Thirteen records specify that their intended audiences are emergency response development and implementation professionals. The remaining four studies do not state an intended audience.^7, 26, 27, 32^ Policymakers and public health officials are the most prevalent of the explicitly mentioned target audiences.

### Behavioural Science Approaches and Theories

The included papers recommend numerous approaches and theories (Table 1). There are multiple uses of the Behaviour Change Wheel (BCW),^5, 26-28^ COM-B,^5, 27^ and social identity theory.^24, 29^ Three records that do not utilise theory,^22, 30, 34^ and two rely on behavioural science constructs instead.^25, 32^

The two records discussing decision-making tools do not utilise behavioural science theory.^22, 34^ The third record that does not use theory is a narrative review of a selection of protective behaviours.^30^

Three records recommend a multidisciplinary approach to behaviour change interventions in an emergency response including stakeholders from various backgrounds to aid in the translation of specialist knowledge into lay terms so that it can be rapidly utilised by emergency response practitioners.^7, 23, 26^

### Validation of tools

Four records state that their tool has been tested or validated.^5, 24, 30, 31^ Six records involve established models, such as the BCW and COM-B, that have been validated previously.^5, 23,25-28^ Morton et al. (2021) draw on the Person-Based Approach, which has been validated; however, they do not mention whether their adapted Table of Changes has been tested. ^7^ Similarly, based on what is said in the paper, it is unclear whether the Template for Rapid Iterative Consensus of Experts (TRICE) framework, which draws on the BCW and COM-B, has been tested.^5^ Linnemayr et al. propose behavioural economics methods that have been validated in other fields but not in emergency response.^25^ Three of the records that do not mention testing or validation cite a scientific evidence base that forms the basis of their tool,^26, 29, 33^ suggesting that core concepts have been tested even if the tool itself has not yet. The two records that do not come from academic literature do not state whether they have been tested or validated.^20, 21^

### The ACE criteria

This section considers how the content of the included records relates to the ACE criteria outlined in the introduction. Five records discuss co-production in developing or implementing an intervention.^7, 21, 23, 26, 32^ They do so by recommending the involvement of stakeholders in the intervention design,^7, 21^ recommending using community figures to implement bottom-up intervention,^32^ and co-production with the target population.^23, 26^ The remaining studies do not mention co-production. There is a range within the included records in the extent to which the tools consider how to engage and support people from diverse contexts and underserved communities. Five records have this consideration as a part of the tool by recommending participation of local stakeholders to aid in developing tailored interventions and how to gain community support.^23, 26, 29, 30, 34^ For example, Jackson et al. identified that engaging with pre-existing community organisations and community leaders improved the delivery of rapid behavioural science interventions by making communities feel involved in the response.^23^ Two records do not have it as part of the tool but do discuss targeted messaging and how to reach isolated communities.^21, 25^ Of the records that do not discuss engagement with people from diverse contexts, Michie et al. (2020) mention that equity concerns require further research, and two others discuss models which allow for consideration of diverse or underserved communities.^32, 33^ Only three records incorporate co-production with the target population and consider engagement with diverse communities.^21, 23, 26^

Four records incorporated rapid testing, evaluation or validation methods for interventions,^5, 7, 27, 28^ two of which used the Acceptability, Practicability, Effectiveness, Affordability, Spill-over effects, and Equity (APEASE) criteria.^27, 28^ A further two records discuss testing, evaluation or validation methods for interventions, but do not mention if it is designed to be used rapidly.^21, 26^ Two of the three records that are reviews identify that little of the literature they reviewed included testing, evaluation or validation of interventions and that future work needs to focus on methodically incorporating this into all interventions.^23, 33^

Six records clearly state that their tool is designed to be implemented rapidly.^5, 7, 20, 22, 28, 30^ Chater et al. recommend doing so using their template to aid in quicky collating scientific evidence into a single message and translating it into language that can be understood by those implementing it.^5^ The remaining records do not state a timeframe for intervention development or implementation using their tool.

None of the seventeen included records meets all of the ACE criteria, and only three meet two of the three criteria.^5, 7, 28^ Seven records meet one criteria,^20-23, 26, 27, 30^ and seven records do not explicitly meet any of the criteria^24, 25, 29, 31-34^ (Figure 3). Three records included co-production with consideration of diversity, six records met the rapid utilisation criterion, and four met the intervention evaluation criterion (Figure 4).

**Figure 3.**
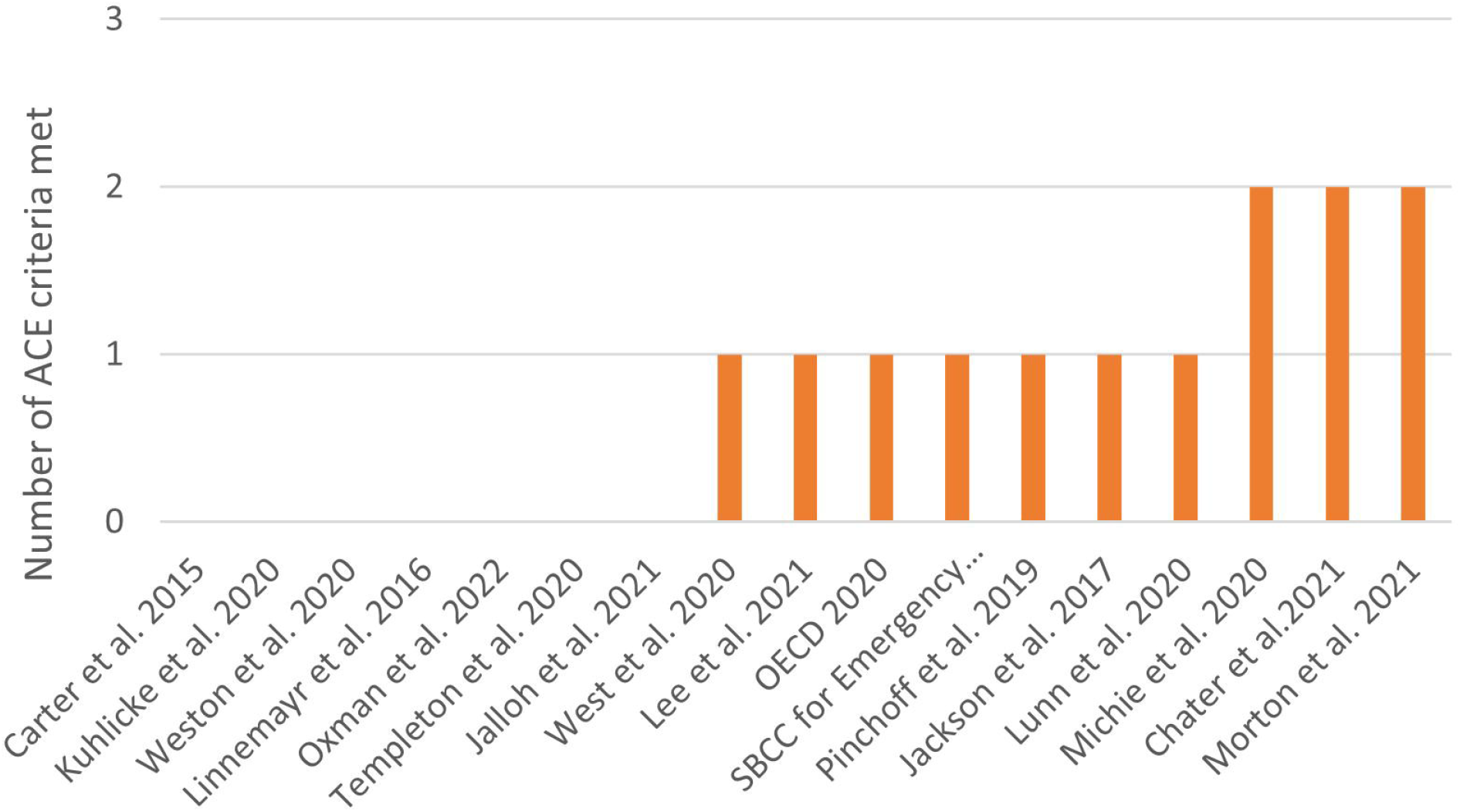
Number of ACE criteria met by the included records

**Figure 4.**
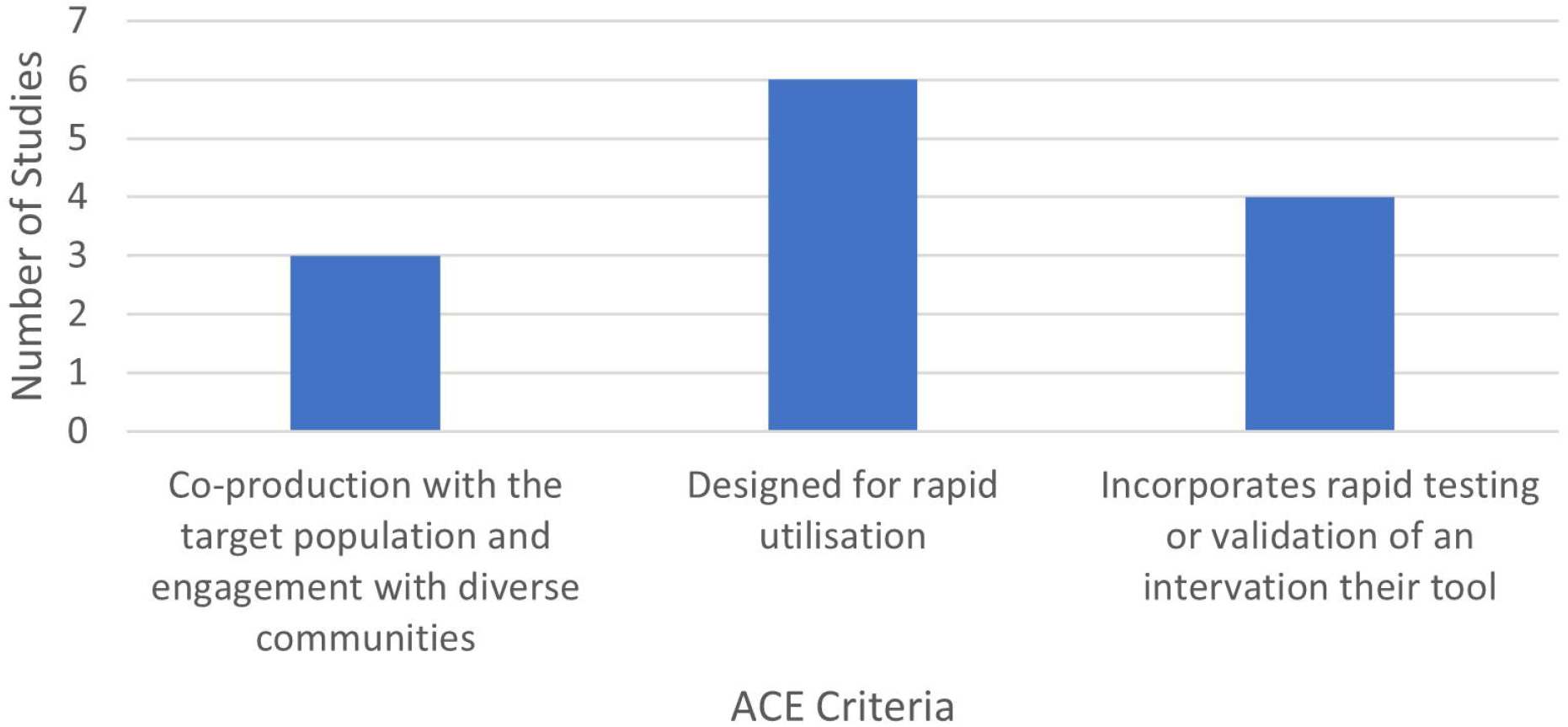
Number of Studies to meet each ACE criterion

## Summary

Commonalities between the included studies are:

- The challenge of identifying behavioural science knowledge in an emergency context, and the importance of consensus so this knowledge is communicated in a united message. ^5,20,22,28^
- The importance of a multidisciplinary approach to behaviour change interventions in emergency response.^7, 23, 26^
- A lack of robust evaluation of past applications of behavioural science in emergency contexts has left practitioners with a limited evidence base.^7, 22, 26, 27^

## Discussion

Overall, the findings from this review identified that there are relatively few accessible tools designed to facilitate the use and evaluation of behavioural science in intervention and guidance development during emergency responses. Given the increased emphasis on behavioural science in emergency response (exemplified by the increase in frameworks and approaches identified herein that were developed post 2020), it is clearly more important than ever to have flexible tools designed to facilitate the incorporation of behavioural science within incident response.

Of the tools that do exist, none of them incorporate all the components of the recently developed ACE framework.^14^ The ACE framework was devised in recognition of the need for a unified framework that incorporates rapid implementation, co-production with the target population including seldom heard voices from diverse communities, and evaluation of interventions and messages. Their criteria have broadly only been discussed in isolation, but a framework that unifies them is required.^14^ Our results support the need for just such a unified framework, as none of the resources included in this review incorporated all of the a-priori identified ACE criteria, and existence of this gap, with ten of the included studies meeting only one or even none.

When considering the broader literature, there is clear evidence for the importance of the criteria against which we have judged the tools identified within this review for effective emergency response. For instance, co-production is valuable in utilising behavioural science in emergency contexts because members of a target community contribute knowledge, skills and access to networks that can enhance an intervention’s effectiveness.^35-37^ Similarly, the COVID-19 pandemic and consequent widening of health inequalities demonstrate that considering how to engage diverse populations is critical in emergency response, and behavioural science can be vital to understanding how.^13, 35, 37, 38^ Consideration of diverse, minority and disproportionately affected populations can identify and overcome barriers to engagement with health services, tackle stigma, increase resilience, and empower communities, all of which are crucial for emergency response and recovery.^35, 39-41^

Furthermore, in terms of a theoretical basis for these tools, the most prevalent approaches in the literature were the BCW, COM-B and social identity theory. BCW and COM-B are appropriate for emergency response because they are designed to be adaptable to various target behaviours and contexts.^26, 42^ Social identity theory can be valuable to emergency response as it can enhance the target population’s ability and willingness to adhere to guidance.^24, 29^ The most commonly used approaches to behavioural science therefore seem appropriate to the emergency response context

In line with the ACE framework, we therefore recommend that future tools cover theory-driven and evidence-based rapid implementation, co-production with the target population, consideration of engaging under-served communities, and rapid evaluation of interventions. Specific attention should be given to covering all the criteria together to maximise the potential impact of future interventions, messaging and guidance during an emergency.

Finally, it is also worth noting that the tools identified within this review are generalised in whom they are intended to be used by. No records are explicitly directed towards specific organisations; however, they all appear to relate to the central management of an emergency; this implies that they were written with central government in mind. How central and local governments and external stakeholders can utilise behavioural science differ, as is demonstrated by the separate guidance documents for use in non-emergency situations.^4, 12^ It is vital for future emergency preparedness that stakeholders can access behavioural science guidance that addresses their capacities and opportunities for implementation, as the UK government’s Emergency response and recovery guidance demonstrates by breaking down the roles of different stakeholders.^43^ Therefore, we recommend that the intended audiences and the needs of said audience be carefully considered when developing such frameworks and toolkits in future.

### Strengths

We are confident in our findings’ internal validity due to the research’s methodological strengths – we prepared a review protocol, searched multiple databases to reduce selection bias, captured grey and published literature, and screened records in duplicate to minimise human error.

### Limitations

Resource limitations meant we could not include potentially relevant hand-identified records in our synthesis or perform data extraction in duplicate.

We have included a list of some of the hand-identified records in Supplementary Material 2, as they may add valuable insight for future work in this topic area. These records were primarily identified after the literature searches had been conducted.

## Conclusions

Behavioural science has significant potential in public health emergency responses. However, it is challenging to implement rigorously and rapidly, which makes having pre-existing tools to guide rapid implementation critical.

Future work into this topic area needs to focus on how to create frameworks that utilise all ACE criteria; are adaptable to a range of potential future public health emergencies; have robust methods for rapid knowledge translation; prioritise multidisciplinary approaches; and include plans to evaluate the intervention in order to strengthen the evidence-base. This research is in line with ACE’s aim of producing an effective evidence-based framework for utilising behavioural science in response to a public health emergency.

## Supporting information

Supplementary Material 1: Search Strategies

Supplementary Material 2: Hand-identified relevant records

## Data Availability

All data produced are available online at the locations provided in the reference list.

## Acknowledgements

AZM conducted the research as an MSc dissertation for the University of Bristol. DW, CF and JK supervised the dissertation and participated in the preparation of the manuscript for publication.

