## Supplementary Material 1: Search Strategies for "A scoping review of behavioural science approaches and frameworks for health protection and emergency response"

MEDLINE and EMBASE search strategy

1. behavioral sciences/ or behavioral research/ or psychology/ or social sciences/
2. Health Behavior/
3. (behavio* science* or science*, behavio*).ti,ab,kw.
4. (behavio* change or change, behavio*).ti,ab,kw.
5. (behavio*, health or behavio*, health-related or health behavio* or health related behavio* or health-related behavio*).ti,ab,kw.
6. (campaign*, health or health campaign* or health promotion* or promotion*, health).ti,ab,kw.
7. Adaptive behavior/
8. 1 OR 2 OR 3 OR 4 OR 5 OR 6 OR 7
9. Emergencies/
10. disasters/ or disaster planning/ or emergencies/ or mass casualty incidents/ or natural disasters/
11. (emergenc* respon* or emergenc* health prepar* or emergenc* resilien*).ti,ab,kw.
12. (disaster planning* or disaster relief planning* or planning*, disaster or planning*, disaster relief or relief planning*, disaster).ti,ab,kw.
13. (crisis intervention* or critical incident stress debriefing or intervention*, crisis).ti,ab,kw.
14. (EPRR or "Emergency Preparedness, Resilience and Response").mp.
15. (rapid respons* or rapid implement*).ti,ab,kw.
16. 9 OR 10 OR 11 OR 12 OR 13 OR 14 OR 15
17. 8 AND 16
18. Limit 17 to yr=”2014-Current”

PsychINFO search strategy

1. behavioral sciences/ or social sciences/ or psychology/ or behavior/ or sociology/
2. health behavior/ or behavior/ or preventive health behavior/ or health knowledge/ or health promotion/ or public health campaigns/
3. (behavio* science* or science*, behavio*).ti,ab.
4. (behavio* change or change, behavio*).ti,ab.
5. (behavio*, health or behavio*, health-related or health behavio* or health related behavio* or health-related behavio*).ti,ab.
6. (campaign*, health or health campaign* or health promotion* or promotion*, health).ti,ab.
7. 1 OR 2 OR OR 3 OR 4 OR 5 OR 6
8. emergency preparedness/ or disasters/ or natural disasters/ or terrorism/
9. (emergenc* respon* or emergenc* health prepar* or emergenc* resilien*).ti,ab.
10. (disaster planning* or disaster relief planning* or planning*, disaster or planning*, disaster relief or relief planning*, disaster).ti,ab.
11. (crisis intervention* or critical incident stress debriefing or intervention*, crisis).ti,ab.
12. (EPRR or "Emergency Preparedness, Resilience and Response").mp.
13. (rapid respons* or rapid implement*).ti,ab.
14. 8 OR 9 OR 10 OR 11 OR 12 OR 13
15. 7 AND 14
16. limit 15 to yr="2014 -Current"

Google search

1. “Behavioural insights, emergency response, EPRR, disaster planning, health protection, behavioural science”
