## Supplementary Material 2: Hand-identified relevant records for "A scoping review of behavioural science approaches and frameworks for health protection and emergency response"

1. Bonell C, Michie S, Reicher S, West R, Bear L, Yardley L, et al. Harnessing behavioural science in public health campaigns to maintain ‘social distancing’ in response to the COVID-19 pandemic: key principles. J Epidemiol Community Health. 2020 74(8):617-9.
2. British Psychological Society. Behavioural science and disease prevention: Psychological guidance. 2020. Available from: <https://www.bps.org.uk/sites/www.bps.org.uk/files/Policy/Policy%20-%20Files/Behavioural%20science%20and%20disease%20prevention%20-%20Psychological%20guidance%20for%20optimising%20policies%20and%20communication.pdf>
3. Corker E, Altieri E, Michie S. Enabling countries to apply behavioural science in using global survey data to inform their Covid-19 policies. Qeios. 2021.
4. Collective Service. COVID-19 Behaviour Change Framework. 2021. Available from: <https://www.rcce-collective.net/wp-content/uploads/2021/03/RCCE-COVID-19-Behaviour-Change-Framework.pdf>.
5. López Gómez A, Dogmanas D, Brunet-Adami N, Bagattini N, Bernardi R. Using behavioural and social sciences to inform public policies during COVID-19, Uruguay. Bull World Health Organ. 2021 99(11):843-4.
6. Michie S. Behavioural strategies for reducing covid-19 transmission in the general population. 2020. Available from: <https://blogs.bmj.com/bmj/2020/03/03/behavioural-strategies-for-reducing-covid-19-transmission-in-the-general-population/>.
7. SAGE. Using behavioural science to help minimise the spread of Covid-19. 2021. Available from: <https://www.independentsage.org/wp-content/uploads/2021/11/12th-November-Behavioural-Science-report.pdf>.
8. Tanis C, Nauta F, Boersma M, van der Steenhoven M, Borsboom D, Blanken T. Practical behavioural solutions to COVID-19: Changing the role of behavioural science in crises. Unpublished. DOI: 10.31234/osf.io/q349k.
9. UCL Centre for Behaviour Change. Responding to COVID-19: contributions from the Centre for Behaviour Change. 2020. Available from: <https://blogs.ucl.ac.uk/cbc-covid/>.
10. World Health Organisation. Communicating risk in public health emergencies: A WHO guideline for emergency risk communication (ERC) policy and practice. 2017. Available from: <https://www.who.int/emergencies/risk-communications>.
11. Williams S, Drury J, Michie S, Stokoe E. Covid-19: What we have learnt from behavioural science during the pandemic so far that can help prepare us for the future. 2021. Available from: <https://www.bmj.com/content/375/bmj.n3028.short>.
